# A year in the public life of COVID-19 and Vitamin D: Representation in UK news media and implications for health communications

**DOI:** 10.1101/2022.04.13.22273832

**Authors:** Alexandra Mavroeidi, Ryan Innes, Esperanza Miyake, Diane Pennington

## Abstract

**Objectives:** To investigate how the relationship between vitamin D and COVID-19 has been presented in traditional media sources (UK newspapers) and assess the level of misinformation associated with this issue by comparing newspaper article content to the evidence-based guidelines.

**Design:** Qualitative study

**Setting:** Data were collected over the first year of the pandemic (February 2019-20), from the five most widely read UK based newspapers by searching the Nexis database using keywords “vitamin D” and “COVID”. An inductive thematic analysis was carried out on the data to identify themes and subthemes. Quality control of the coding was conducted on a sample of the dataset (20%). Data were also compared to the “ground truth” identified as the NICE report titled “COVID-19 rapid guideline: vitamin D” to explore the accuracy of media outputs.

**Results:** The four main themes identified were ‘association of vitamin D with COVID-19’, ‘politically informed views’, ‘vitamin D deficiency’ and ‘vitamin D sources’. When compared to the ground truth, most of the information from newspaper articles relating to the key findings was ‘correct’ for each of the findings, except for COVID-19 infection.

**Conclusions:** Although most of the information included in newspaper articles concerning the relationship of vitamin D with COVID-19 was ‘correct’, this study highlighted that there was still a notable amount of ‘incorrect’ information published. In the context of COVID-19, it is imperative that media outputs are accurate and inform the public and front-line health professionals as correct information is a key factor in disease prevention. Future research should focus on the accuracy of media outputs to further investigate health misinformation as an issue in traditional media and how that may affect public health. Attempts should be made to improve journalistic integrity through more rigorous and standardised regulations enforced across all media outlets so that public knowledge on current events is based on evidence rather than conjecture.

**Summary boxes:** 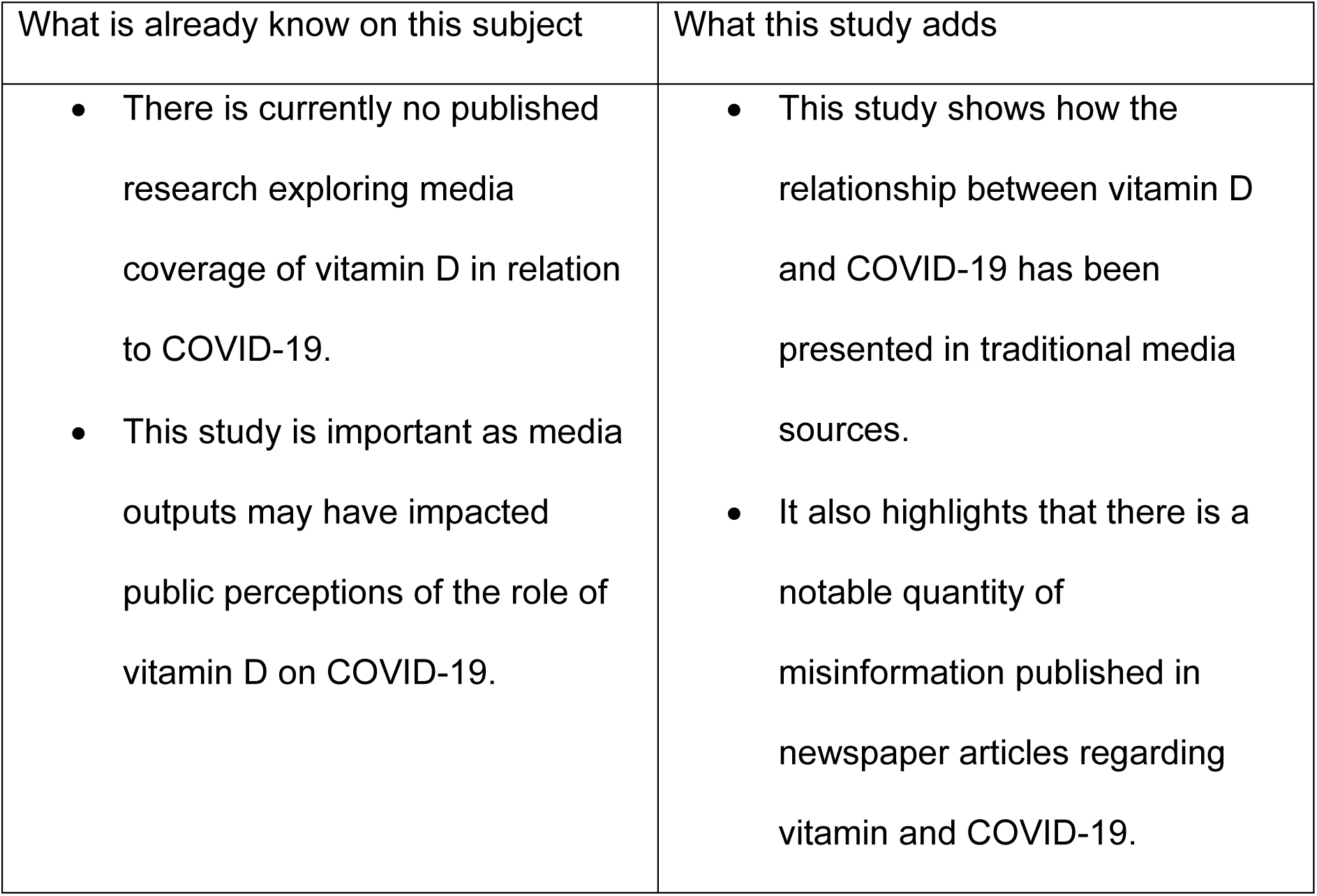

**Data sharing statement:** We are happy to share any data if/when asked.

## Introduction

The current COVID-19 pandemic caused by the severe acute respiratory syndrome coronavirus 2 (SARS-CoV-2) is a subject of global concern.^[1]^ There has been an unprecedented response across several sectors to develop targeted therapeutics in an attempt to slow the rate of infection.^[2]^ One modifiable lifestyle intervention that has received particular interest is dietary vitamin D.^[3]^

The scientific debate basis for the potential relationship of vitamin D status with COVID-19 is based upon the association of low serum concentration of 25(OH)D with increased susceptibility to acute respiratory tract infections.^[4]^ Furthermore, antimicrobial effector mechanisms such as induction of autophagy and synthesis of reactive nitrogen intermediates and reactive oxygen intermediates are reportedly initiated by vitamin D metabolites.^[5]^ This suggests that vitamin D may provide the host with protection against respiratory pathogens, effectively reducing their risk of contracting an acute respiratory tract infection. It is theorized that, this may translate to SARS-CoV-2 infection. Several correlational studies have reported a relationship between vitamin D deficiency and poor COVID-19 outcomes.^[4,6,7]^ However, a number of risk factors for poor COVID-19 outcomes are similar to those for vitamin D deficiency such as old age and darker skin.^[8,9]^ Consequently, there are several other risk factors that must be considered before the true extent of the cause-and-effect relationship between vitamin D and COVID-19 can be extrapolated. This was highlighted in the evidence based rapid review by the National Institute for Health and Care Excellence (NICE), in collaboration with Public Health England and the Scientific Advisory Committee on Nutrition.^[10]^. Their recommendations supported the current government advice, urging everyone to take vitamin D supplements to maintain bone and muscle health (autumn and winter months) in the UK. They were also in line with the new guidance from the UK government^[11]^, which highlighted that extremely clinically vulnerable people can opt in to receive a free 4-month supply of daily vitamin D supplements.

Previous research has highlighted the vital role that the media plays in the distribution of information to the public.^[12,13]^ Individuals become particularly dependant on media outputs for information when they don’t have direct knowledge or personal experience of a subject. As COVID-19 is a current pandemic, the level of direct knowledge and personal experience is relatively low. Therefore, the public are especially dependant on the media for information concerning COVID-19.^[14-16]^.

The aim of this study is to investigate how the relationship between vitamin D and COVID-19 has been presented in traditional media sources (UK newspapers). In addition we aimed to assess the level of misinformation associated with this issue by comparing newspaper article content to the evidence-based guidelines from the NICE report.

## Methods

### Data sources and Collection

Data were collected from the five most popular newspapers (The Mirror, The Sun, The Daily Mail and Mail on Sunday, Metro and Express Online) published in the UK as ranked by monthly reach from April 2019 to March 2020.^17^ Data were collected using Nexis; a database of media outputs that was accessed through the Strathclyde Library. The keywords “covid” and “vitamin D” were used to search on Nexis on 19/2/2021.

The Nexis search was carried out in April 2021, and was limited to Newspaper Articles published over a year period (February 2020 to February 2021). These provided data on the number of articles published each month containing the keywords throughout the pandemic (Figure 1). Figure 2 shows the number of articles published in each of the five most popular newspapers that were included in the analysis. The results of the search were imported to NVivo 12 for analysis.

**Figure 1.**
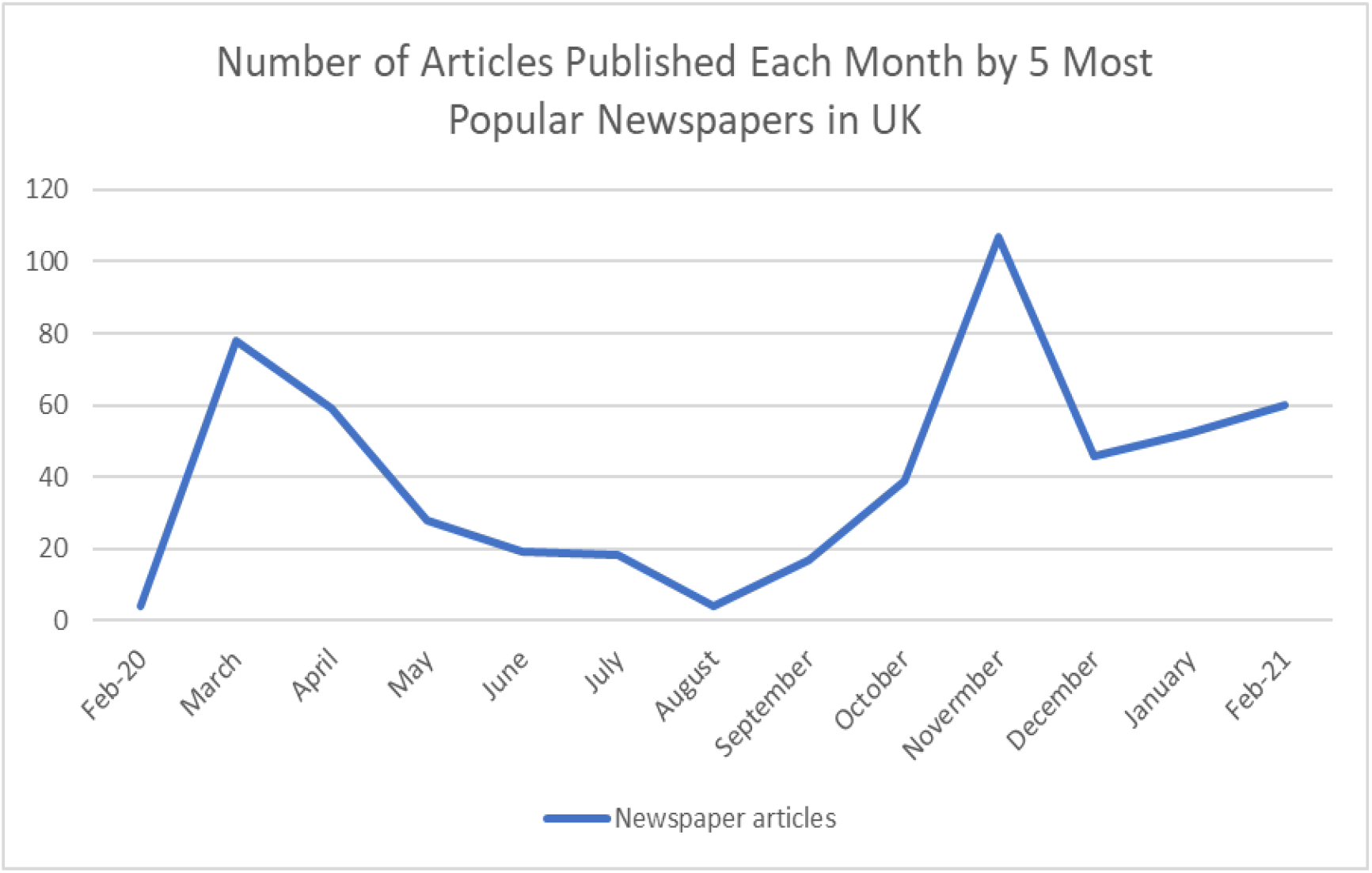
Number of Articles published by the five most popular newspapers in the UK containing the keywords “vitamin D” and “COVID” over 1 year of publications. Retrieved 13/4/21.

**Figure 2.**
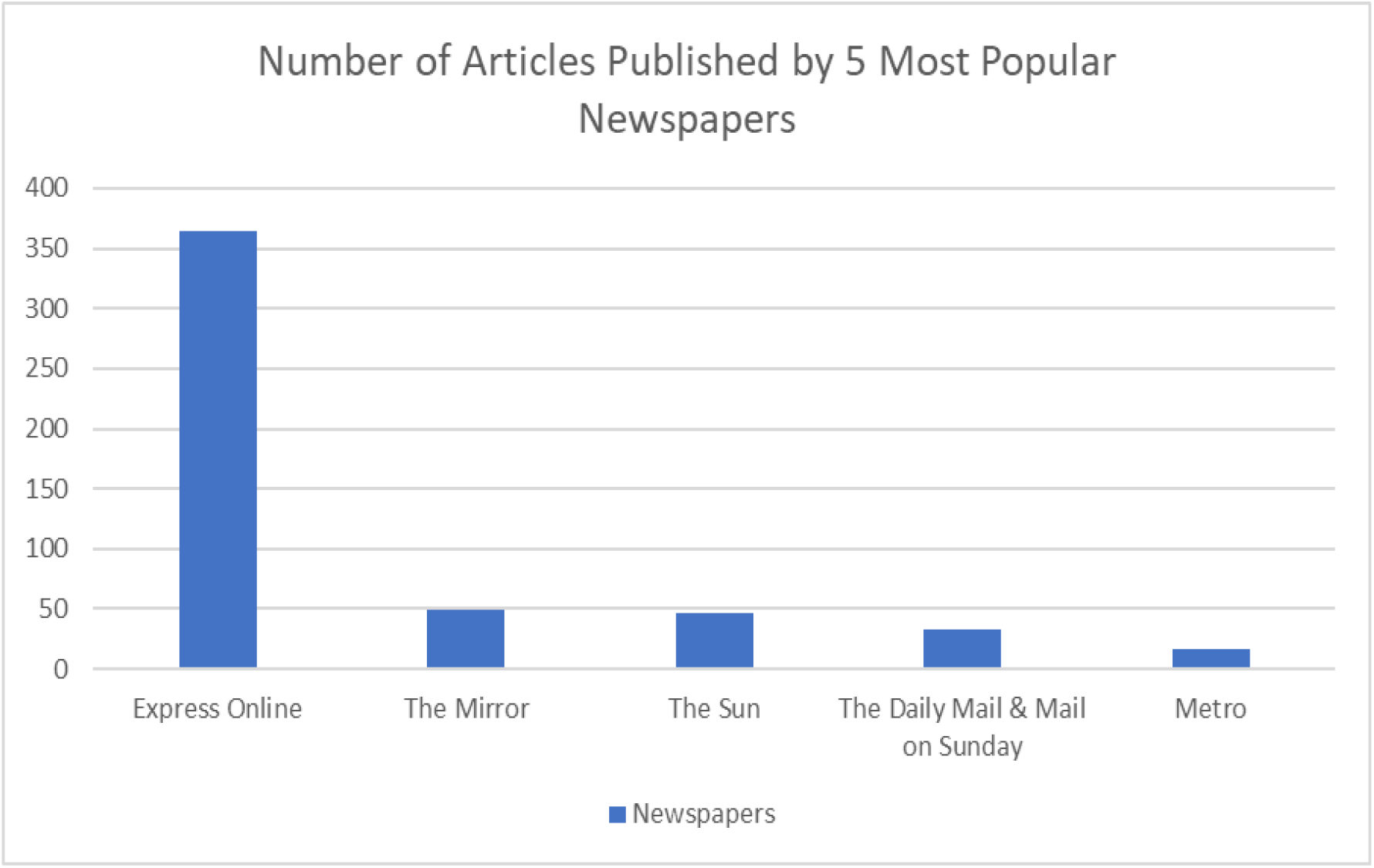
Number of Articles published in each of the five most popular newspapers in the UK containing the keywords “vitamin D” and “COVID” over 1 year of publications. Retrieved 13/4/21.

### Data Analysis

Inductive thematic analysis was used to investigate how the relationship between vitamin D and COVID-19 has been presented in newspaper articles. Thematic analysis is a qualitative technique that enables researchers to extract meanings and concepts from data sets that cannot be quantitively analysed, such as focus group transcripts ^18^. The technique is commonly utilized in health settings to analyse patient experiences ^[19-21]^ and is often deployed in social media based research ^[22-24]^. An inductive approach was used, meaning that the analysis was data driven rather than using a pre-determined coding frame ^[25]^. Therefore, the themes identified are strongly linked to the data, as data were coded without trying to fit them into a pre-existing coding frame ^[25]^. The step-by-step guide outlined in (Braun & Clarke, 2006) was utilised to analyse the data. In short the six steps followed were: familiarisation data, generating initial codes, searching for themes, reviewing themes, defining and naming themes, and producing the report. The inductive thematic analysis was carried out in NVivo 12 (Appendix A).

Quality control of the coding was conducted by another independent researcher with previous experience of thematic analysis (BH), who coded a sample of the dataset (20%). The researcher who carried out the quality control (BH) agreed with the provisional themes and sub themes identified by the main researcher (RI).

The codes identified from analysis of the newspaper articles were also compared to the “ground truth” in order to explore the accuracy of media outputs. The ground truth “refers to an external method to assess the truth or credibility of messages” ^[26]^. The NICE report titled “COVID-19 rapid guideline: vitamin D” was used as the ‘ground truth’ because it summarises evidence-based recommendations developed by an independent committee ^[10]^. The four key findings identified from the NICE report were:

1. Do not offer a vitamin D supplement to people solely to treat COVID-19.
2. Do not offer a vitamin D supplement to people solely to prevent COVID-19.
3. Low vitamin D status was associated with more severe outcomes from COVID-19. However, it is not possible to confirm causality because many of the risk factors for severe COVID-19 outcomes are the same as the risk factors for low vitamin D status.
4. Adults, young people and children over 4 years should consider taking a daily supplement containing 10 micrograms of vitamin D between October and early March. Groups at high risk of vitamin D deficiency should also consider taking a daily supplement containing 10 micrograms of vitamin D year-round.

Data that did not relate to the key findings outlined above were excluded from this portion of the analysis. Appropriate data were compared to the key findings and coded in NVivo into either ‘correct’ or ‘incorrect’ nodes.

## Results

### Descriptive

The number of newspaper articles published by the five most popular newspapers in the UK containing the keywords “vitamin D” and “COVID” declined from March to August before rising again, peaking in November (Figure 1).

The majority of newspaper articles collected from the keyword search conducted on Nexis on 19/2/2021 were published in Express Online. Comparatively, each of the four other newspaper sources published very few articles containing the keywords “vitamin D” and “COVID” (Figure 2).

When looking at the data from the five newspaper sources (The Mirror, The Sun, The Daily Mail and Mail on Sunday, Metro and Express Online) four main themes and twelve sub-themes were identified (Table 1, Appendix A & B).

**Table 1.** Summary of themes and subthemes for newspaper article data.

| Theme | Subtheme |
| --- | --- |
| Association of Vitamin D with COVID-19 | COVID-19 Infection<br>COVID-19 Outcomes<br>COVID-19 Treatment |
| Politically Informed Views | MPs<br>Government Measures<br>Vitamin D Guidelines |
| Vitamin D Deficiency | High Risk Groups<br>Impact of Vitamin D on Health<br>Prevalence of Vitamin D Deficiency |
| Vitamin D Sources | Food<br>Sunlight<br>Supplementation |

### Theme 1: Association of Vitamin D with COVID-19

#### Subtheme 1A: COVID-19 Infection

Vitamin D was commonly associated with risk of COVID-19 infection, with several articles quoting ‘scientists’ as their source.

> *“VITAMIN D: A lack of this vitamin is associated with increased risk of coronavirus infection.” (The Sun 1)*
>
> “*Scientists have claimed that vitamin D could lower your risk of becoming infected with coronavirus.” (Express Online 1)*

However, some articles did highlight the importance of following government guidelines that aim to reduce risk of COVID-19 infection.

> *“Anyone on social media trying to distract from the established message of handwashing, alcohol gel and social distancing is putting people’s lives at risk.” (The Daily Mail 1)*

### Subtheme 1B COVID-19 Outcomes

The majority of media coverage regarding the potential relationship of vitamin D with COVID-19 outcomes focussed on findings from published research, resulting in several different figures being shared. Additionally, the studies were not fully referenced in the majority of articles.

> *“Vitamin D reduces COVID-19 deaths by an astounding 60 percent, a study has found.” (Express Online 2)*
>
> *“The research found Covid patients who were given doses of vitamin D were 80 percent less likely to require ICU treatment.” (Express Online 3)*

Some articles inferred a causal link between vitamin D and COVID-19 outcomes, whereas others highlighted the need for further research.

> *“This doesn’t prevent you from getting COVID-19, but certainly there’s a lot of evidence to show that it reduces the severity of the illness.” (Express Online 4)*
>
> *“While a number of studies have suggested that the vitamin may also reduce mortality from Covid, reviews by health officials have suggested there is insufficient evidence.” (Express Online 5)*

### Subtheme 1C: COVID-19 Treatment

Opinions shared on the use of vitamin D as a COVID-19 treatment varied. Some supported the use of vitamin D as a treatment, while others objected.

> *“Dr Hernández commented on the findings: “Vitamin D treatment should be recommended in COVID-19 patients.” (Express Online 6)*
>
> *“VITAMIN D does not help to prevent or treat coronavirus, health chiefs insisted after a review.” (The Sun 2)*

Cases where vitamin D was used as treatment in COVID-19 patients were also shared.

> *“Donald Trump is also being treated with a cutting-edge antibody drug - the anti-viral drug remdesivir - along with vitamin D, melatonin, zinc, an antacid called famotidine and aspirin.” (The Daily Mail 2)*
>
> *“He added: “In defiance of guidelines from the HSE, I use hydroxychloroquine, azithromycin, and readily available vitamin D on high risk patients who got sick with Covid in my patient population.” (The Sun 3)*

### Theme 2: Politically Informed Views

#### Subtheme 2A: MPs

Claims made by MPs were reported regardless of their accuracy. David Davis received particular attention as he continually supported widespread supplementation.

> *“Mr Davis said on Twitter: “The findings of this large and well conducted study should result in this therapy being administered to every Covid patient in every hospital in the temperate latitudes. Furthermore, since the study demonstrates that the clear relationship between vitamin D and Covid mortality is causal, the UK government should increase the dose and availability of free vitamin D to all the vulnerable groups.” (Express Online 7)*
>
> *“Tory backbencher David Davis and Labour MP Rupa Huq have been leading the charge in raising awareness of the supposed effects of the vitamin on Covid-19 in the UK. Mr Davis said: “All the observational studies show strong vitamin D effects on infectiousness, morbidity and mortality.” (Express Online 8)*

#### Subtheme 2B: Government Measures

It was theorized that vitamin D levels of the population would fall due to extended periods of lockdown.

> *“Due to months of lockdown a lot of people have also been spending the majority of their time indoors over the last year, reducing their chances to get enough vitamin D from the sun.” (Express Online 9)*
>
> *“One in five people in the UK, under normal circumstances, has a vitamin D deficiency. Now, with the coronavirus lockdown meaning little time outside in sunlight, our levels could be even lower.” (The Daily Mail 3)*

#### Subtheme 2C: Vitamin D Guidelines

Several articles accurately detailed the recommended daily dose of vitamin D.

> *“Vitamin D UK RDA = 10mcg a day for men and women” (Express Online 10) “Most people need around 10mcg of vitamin D a day, including pregnant and breastfeeding women. Any babies younger than one year old need between 8.5 and 10mcg daily.” (Express Online 11)*

However, there was one instance where the conversion from micrograms to International Units was reported incorrectly.

> *“The equivalent of 10mcg in IU is 40IU.” (Express Online 12)*

### Theme 3: Vitamin D Deficiency

#### Subtheme 3A: High Risk Groups

Particular groups were reported of being at higher risk of vitamin D deficiency due to a number of factors.

> *““You may find you’re at an even higher risk if you have a darker skin type, are older, or regularly wear clothing that covers your skin. Certain health conditions can leave you vulnerable to low vitamin D levels too, such as having a BMI that falls in the obese category, having kidney or liver disease, or if you take anti-epilepsy medications.” (Express Online 13)*

It was often recommended that those in high-risk groups consider supplementation to reduce the likelihood of becoming vitamin D deficient.

> *“Ethnic minority groups with dark skin should also consider taking a year-round supplement.” (The Daily Mail 4)*
>
> *“Some groups who don’t tend to get much sun exposure even in the summer, such as those in care homes, as well as some with darker skins who need more sun exposure to produce enough vitamin D, should take a supplement all year round.” (The Daily Mail 5)*

#### Subtheme 3B: Impact of Vitamin D on Health

Regardless of its potential association with COVID-19, vitamin D was reported of playing several important roles in the maintenance of general health.

> *“Asked why vitamin D is so important for the body, Dr Deo said: “Vitamin D is essential for the human body to work as it should. It regulates the amount of calcium and phosphate in your body - nutrients which are needed for healthy teeth, bones and muscles. As well as this, vitamin D helps our muscles to stay healthy and in good working order. The immune system also uses it to fight off bacteria and viruses, helping us to stay healthy.” (Express Online 14)*

The association of vitamin D with respiratory tract infections was also commonly discussed and linked to COVID-19.

> *“The new Irish research has highlighted the key role vitamin D plays in preventing respiratory infections, which could have important implications for the fight against COVID-19.” (Express Online 15)*
>
> *“Research suggests that low vitamin D levels may increase the risk of respiratory tract infection including COVID19.” (Express Online 16)*

#### Subtheme 3C: Prevalence of Vitamin D Deficiency

It was acknowledged that the prevalence of vitamin D deficiency varies throughout the course of each year depending on season, making it difficult to determine exactly what percentage of the population are deficient at any time. Despite this, several different figures were presented in the articles.

> *“About 25 percent of Brits are Vitamin D deficient normally.” (Express Online 17)*
>
> *“However, around half of the UK population is deficient in it.” (The Daily Mail 6)*
>
> *“Most of us have Vitamin D deficiency at the moment.” (Express Online 18)*

### Theme 4: Vitamin D Sources

#### Subtheme 4A: Food

It was recognised that naturally occurring dietary sources of vitamin D are limited, making it hard to achieve optimal vitamin D levels through diet alone.

> *“So even a healthy, well-balanced diet that provides all the vitamins and nutrients you need, is unlikely to provide enough vitamin D if you aren’t able to get enough sun.” (Express Online 19)*
>
> *“Since it’s difficult for people to get enough vitamin D from food alone.” (Express Online 20)*

Fortification was often mentioned in articles that discussed dietary sources of vitamin D, even though very few foods are fortified with vitamin D in the UK. Claims were made about the positive impact fortification would have on the nation’s health.

> *““Adding vitamin D to food would reduce deaths and significantly cut NHS costs,” The Guardian reported. A review of existing data estimates that supplementing food with vitamin D would prevent millions and flu cases and possibly save lives.” (Express Online 21)*
>
> *“Dr Davies added: “Finland is the only country with an effective vitamin D food fortification programme, and has had one of the best pandemic responses globally.” (Express Online 22)*

#### Subtheme 4B: Sunlight

Exposure to sunlight is highlighted as the primary source of vitamin D for most people in the UK and worldwide.

> *“Sunlight is the main source of Vitamin D.” (Express Online 23)*
>
> *“In normal circumstances, sunshine, not food, is where most of your vitamin D comes from.” (Express Online 24)*

However, the negative effects of too much sunlight exposure were also explored, such as increased risk of skin cancer.

> *“It’s not possible to overdose on vitamin D by spending too much time in the sunshine, however. If you choose to spend more time in the sun, don’t forget to wear suncream and protect your skin.” (Express Online 25)*

#### Subtheme 4C: Supplementation

Vitamin D supplements were frequently presented as a necessity.

> *“Vitamin D is a vital addition to your supplement routine.” (Express Online 26)*
>
> *“When it comes to supplements, vitamin D is an absolute must.” (Express Online 27)*

Guidelines state that those who choose to use a supplement should do so with a dose of 10 micrograms (National Institute for Health and Care Excellence, 2020). Despite this, doses higher than 10 micrograms were frequently presented.

> *“So I’d recommend an oral supplement of 1,000IU to 2,000IU daily for most people.” (The Daily Mail 7)*
>
> *“Dr Davies assures “4,000IU daily is perfectly safe and suited to almost all adults”, only advising children and underweight adults to take less.” (Express Online 28)*

However, warnings about the potential risks of high dose supplementation were also often discussed.

> *“If you take too many Vitamin D supplements over a long period of time, calcium could build up in the body and cause hypercalcaemia. This can weaken the bones and damage the kidneys and the heart.” (Express Online 29)*

### Ground Truth

Figure 3 shows the number of ‘correct’ and ‘incorrect’ codes identified when data from the Newspaper Articles were compared to the key findings from the NICE report titled “COVID-19 rapid guideline: vitamin D” (National Institute for Health and Care Excellence, 2020). Most of the information relating to the key findings was ‘correct’ for each of the findings, except for COVID-19 infection.

**Figure 3.**
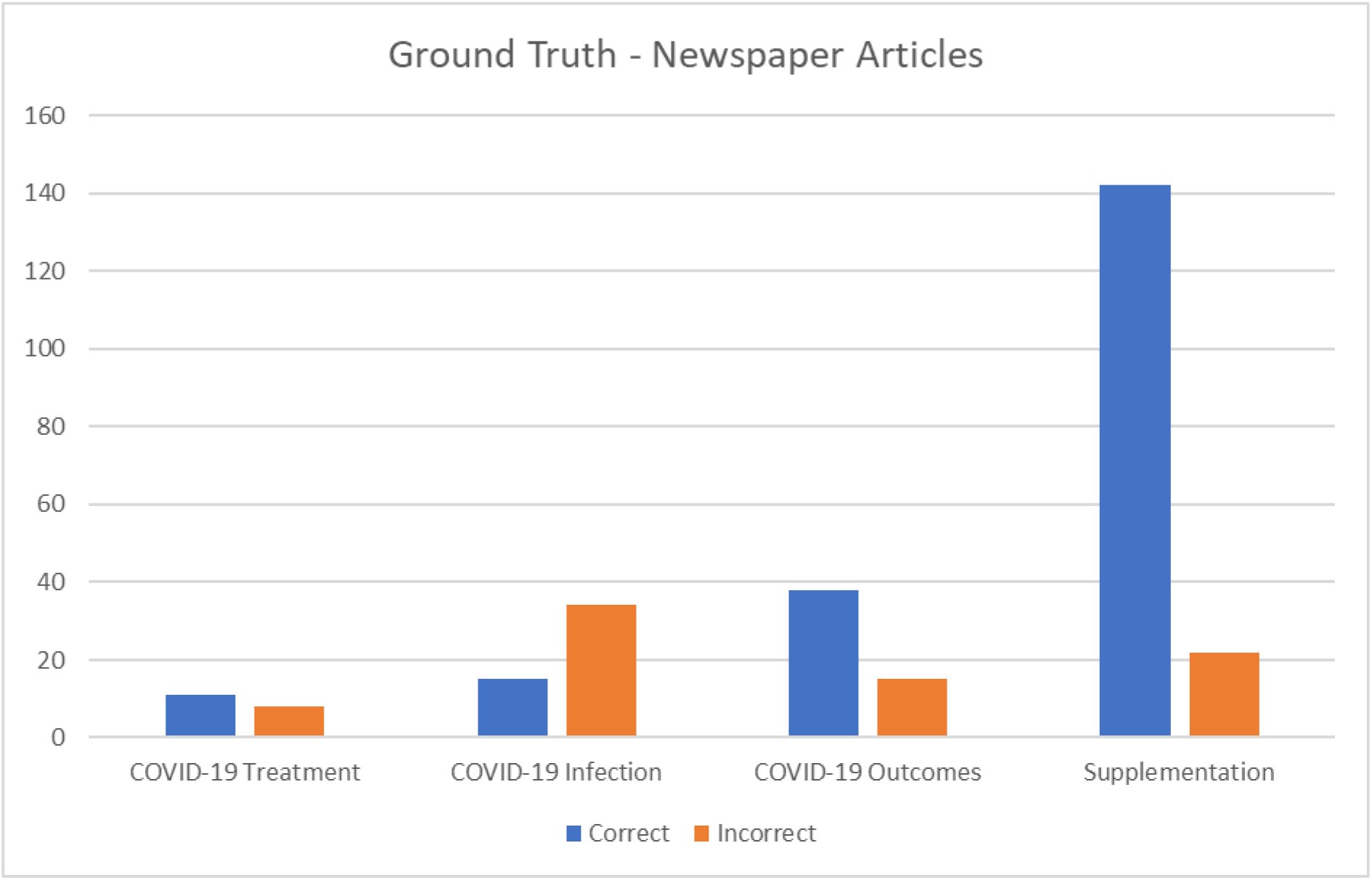
‘Correct’ vs ‘Incorrect’ opinions from newspaper articles.

### 1. COVID-19 Treatment

The NICE report states that vitamin D supplementation should not be used as a COVID-19 treatment. Of the key findings, COVID-19 treatment was the least discussed in the newspaper articles. The majority of codes were ‘correct’.

#### Correct

> *“But there is currently not enough evidence to support taking vitamin D to prevent or treat coronavirus.” (Express Online 30)*

#### Incorrect

> *“Mr Davis called for the therapy to be rolled out in hospitals immediately to “save many thousands of lives”. (Express Online 31)*

### 2. COVID-19 Infection

Vitamin D supplementation should not be offered to prevent COVID-19 infection. This was the only key finding that was reported ‘incorrectly’ more often than ‘correctly’ in the newspaper articles.

#### Correct

> *“There is also no evidence to suggest that vitamin D could help prevent you getting the virus.” (Express Online 32)*

#### Incorrect

> *“But, you could lower your chances of becoming infected with COVID-19 by simply taking vitamin C and D supplements, it’s been claimed.” (Express Online 33)*

### 3. COVID-19 Outcomes

Low vitamin D status is associated with more severe COVID-19 outcomes; however, it is not possible to confirm causality. This was reported ‘correctly’ more than twice as often as ‘incorrectly’.

#### Correct

> *“As COVID-19 is a respiratory illness it may limit the severity of the illness for those who become infected, however there has been no conclusive evidence linking the two.” (Express Online 34)*

#### Incorrect

> *“Furthermore, since the study demonstrates that the clear relationship between vitamin D and Covid mortality is causal, the UK government should increase the dose and availability of free vitamin D to all the vulnerable groups.” (Express Online 7)*

### 4. Supplementation

Adults, young people, and children over 4 years should consider taking a daily supplement containing 10 micrograms of vitamin D between October and early March. Groups at high risk of deficiency should consider year-round supplementation. Supplementation was the most discussed theme. The vast majority of information reported was ‘correct’.

#### Correct

> *“You should take 10 micrograms of vitamin D a day between October and early March.” (Express Online 35)*

#### Incorrect

> *“In light of the current global situation, we recommend taking 5000 IU per day,” recommended Karuzin.” (Express Online 36)*

## Discussion

This study identified how the relationship between vitamin D and COVID-19 has been presented in popular UK based newspapers. The four main themes identified through inductive thematic analysis were association of vitamin D with COVID-19, politically informed views, vitamin D deficiency and vitamin D Sources (Table 1). Generally, when compared to the ground truth, newspaper articles shared a higher proportion of ‘correct’ information than ‘incorrect’ information. However, there was still a notable amount of ‘incorrect’ information published (Figure 3).

There is currently no published research exploring media coverage of vitamin D in relation to COVID-19. However, (Caulfield et al., 2014) examined the nature of media coverage of vitamin D in relation to its role in health and the need for supplements ^[27]^. (Caulfield et al., 2014) collected data from the top five daily newspapers in the UK, USA and Canada. Analysis of popular newspapers from three different countries would have improved the quality of data in this study as it would mean that less of the total data came from Express Online articles. However, (Caulfield et al., 2014) only examined 294 newspaper articles, compared to the 508 newspaper articles analysed in this study. Due to the already high volume of data, it would not have been time feasible to include more newspaper sources.

The strengths and weaknesses of this study must be considered. The collection of articles from the beginning of the pandemic until 19/2/2021 provides a comprehensive view of the nature of UK media coverage regarding the relationship between vitamin D and COVID-19 throughout the course of the pandemic. Also, as the data were collected from the five most popular newspapers, it is fair to assume that these articles would have been the most read in the UK, increasing the likelihood that public opinion was influenced by the articles. However, it must also be noted that the highest frequency of vitamin D and COVID-19 related articles occurred in one newspaper (Express Online). As such a high quantity of the data came from the same source, it is possible that the data was skewed.

Although most of the information included in newspaper articles concerning the relationship of vitamin D with COVID-19 was ‘correct’, this study highlighted that there was still a notable amount of ‘incorrect’ information published. Previous research has indicated that the general population are ill-informed about preventative measures they can take to avoid diseases ^[14]^. In the context of COVID-19, it is imperative that the public are well informed as information is a key factor in disease prevention ^[14]^. Future research should focus on the accuracy of media outputs to explore if misinformation is a widespread issue in traditional media. If misinformation is found to be an issue, attempts should be made to improve journalistic integrity through more rigorous and standardised regulations enforced across all media outlets, so that public knowledge on current events is based on evidence rather than conjecture.

## Data Availability

All data produced in the present study are available upon reasonable request to the authors

## Appendix A Newspaper Articles NVivo Output

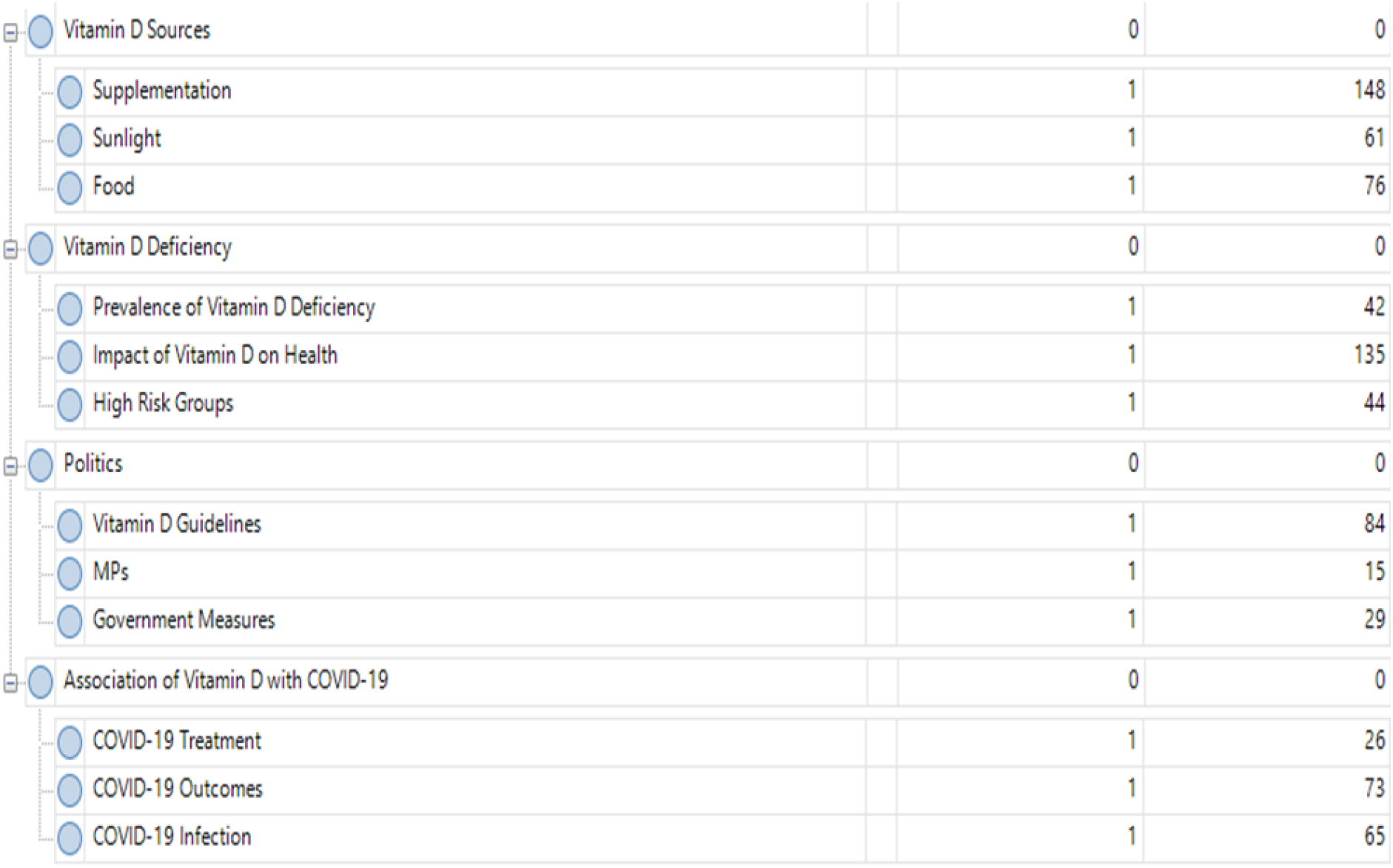

## Appendix B Supplementary Quotes Providing Supportive Evidence for each Theme

Appendix B - Supplementary Quotes Providing Supportive Evidence for each Theme.
| Theme | Subtheme | Quote |
| --- | --- | --- |
| Association of Vitamin D with COVID-19 | COVID-19 Infection | *"When it comes to supplements, vitamin D is an absolute must in order to keep the body operating healthy, but also to help prevent respiratory infections in the case of a coronavirus infection."*<br>*"Studies have found patients with a Vitamin D deficiency are more likely to test positive for the virus."* |
|  | COVID-19 Outcomes | *"This supports the conclusion of a prior pilot trial in Cordoba in which calcifediol treatment led to a reduction of more than 50 per cent of ICU admission in hospitalised Covid-19 patients."*<br>*"As COVID-19 is a respiratory illness it may limit the severity of the illness for those who become infected, however there has been no conclusive evidence linking the two."* |
|  | COVID-19 Treatment | *"Covid treatment: Vitamin D has shown to help reduce serious fatalities by 60 percent."*<br>*"It's thought adequate amounts of vitamin D "might help in the treatment of COVID-19 by preventing the cytokine storm" - a common cause of mortality."* |
| Politically Informed Views | MPs | *"3.18pm update: David Davis presses Matt Hancock on Vitamin D treatment"*<br>*"It comes after Health Secretary Matt Hancock ordered a review into the use of vitamin D to prevent Covid-19."* |
|  | Government Measures | *"About 25 percent of Brits are Vitamin D deficient normally, and the lockdown puts us more at risk of being deficient in Vitamin D"*<br>*"There are concerns among health officials that lockdowns have deprived many of sunshine, which is key to Vitamin D production."* |
|  | Vitamin D Guidelines | *“The Government recommends that everyone should take a daily vitamin D supplement during the winter to avoid a deficiency”*<br>*““You should take 10 micrograms of vitamin D a day between October and early March”* |
| Vitamin D Deficiency | High Risk Groups | *“According to the initiative, deliveries will be made to clinically extremely vulnerable and care home residents. These individuals are at a higher risk of Vitamin D deficiency as these groups have spent more time indoors this year.”*<br>*“We advise that everyone, particularly the elderly, those who don’t get outside and those with dark skin, takes a vitamin D supplement containing 10 micrograms every day.”* |
|  | Impact of Vitamin D on Health | *“Vitamin D is essential when treating acute respiratory infections. Reports obtained from previous studies highlight Vitamin D’s benefit on calcium and bone homeostasis, especially in children, the elderly and osteoporotic patients. In addition, it has a number of other very positive effects on the immune system and lung function which could be crucial in the fight against COVID-19.”*<br>*“A lack of vitamin D can lead to bone deformities such as rickets in children, and bone pain caused by a condition called osteomalacia in adults.”* |
|  | Prevalence of Vitamin D Deficiency | *“Around 20 percent of people in the UK suffer from low vitamin D levels.”*<br>*“Many people are deficient in this vital vitamin”* |
| Vitamin D Sources | Food | *“You can try to make sure your vitamin D levels are high enough by eating foods that are rich in it. “These include cheese, oily fish like mackerel, salmon and tuna, beef liver and egg yolks.”*<br>*“You can top up on Vitamin D by eating specific foods, including oily fish such as* |
|  |  | *salmon, sardines, herring and mackerel, as well as red meat. Liver, egg yolks, and fortified foods such as some fat spreads and breakfast cereals are also good sources of Vitamin D."* |
|  | Sunlight | *"Most people get enough vitamin D during the spring and summer months from sunlight"*<br>*"Direct sunlight encourages the body to naturally produce vitamin D, which is why it's easier for people to make vitamin D in the summer, when it's more inviting outside."* |
|  | Supplementation | *"An easy way to get enough Vitamin D is by taking supplements"*<br>*"people are being encouraged to top up on Vitamin D supplements to make sure they are the healthiest they can be."* |

## References

01 Misra D, Agarwal V, Gasparyan A, Zimba O. Rheumatologists’ perspective on coronavirus disease 19 (COVID-19) and potential therapeutic targets. Clinical Rheumatology. 2020;39(7):2055–2062. doi: 10.1007/s10067-020-05073-9

02 Carter S, Baranauskas M, Fly A. Considerations for Obesity, Vitamin D, and Physical Activity Amid the COVID-19 Pandemic. Obesity. 2020;28(7):1176–1177. doi: 10.1002/oby.22838

03 Lanham-New S, Webb A, Cashman K et al. Vitamin D and SARS-CoV-2 virus/COVID-19 disease. BMJ Nutrition, Prevention & Health 2020;3:106–110. doi:10.1136/bmjnph-2020-000089

04 Ilie P, Stefanescu S, Smith L. The role of vitamin D in the prevention of coronavirus disease 2019 infection and mortality. Aging Clinical and Experimental Research 2020;32:1195–1198. doi:10.1007/s40520-020-01570-8

05 Martineau A, Jolliffe D, Hooper R et al. Vitamin D supplementation to prevent acute respiratory tract infections: systematic review and meta-analysis of individual participant data. BMJ 2017;:i6583. doi:10.1136/bmj.i6583

06 Sulli A, Gotelli E, Casabella A et al. Vitamin D and Lung Outcomes in Elderly COVID-19 Patients. Nutrients 2021;13:717. doi:10.3390/nu13030717

07 Baktash V, Hosack T, Patel N et al. Vitamin D status and outcomes for hospitalised older patients with COVID-19. Postgraduate Medical Journal 2020;:postgradmedj-2020-138712. doi:10.1136/postgradmedj-2020-138712

08 The World Health Organization. Report of the WHO-China Joint Mission on Coronavirus Disease 2019 (COVID-19). 2020. https://www.who.int/docs/default-source/coronaviruse/who-china-joint-mission-on-covid-19-final-report.pdf (accessed 29 Jul 2021).

09 Dey T, Sinha A. Ethnicity and COVID-19 - A commentary on “World Health Oganization declares global emergency: A review of the 2019 novel coronavirus (COVID-19)” (Int J Surg 2020;76:71-6). International Journal of Surgery 2020;83:75–76. doi:10.1016/j.ijsu.2020.08.046

10 National Institute for Health and Care Excellence. COVID-19 rapid guideline: vitamin D. 2020. https://www.nice.org.uk/guidance/ng187/resources/covid19-rapid-guideline-vitamin-d-pdf-66142026720709 (accessed 29 Jul 2021).

11 Vitamin D and clinically extremely vulnerable (CEV) guidance. (2022). Retrieved 13 April 2022, from https://www.gov.uk/government/publications/vitamin-d-for-vulnerable-groups/vitamin-d-and-clinically-extremely-vulnerable-cev-guidance

12 Happer C, Philo G. The Role of the Media in the Construction of Public Belief and Social Change. Journal of Social and Political Psychology 2013;1:321–336. doi:10.5964/jspp.v1i1.96

13 Chapman S, Haynes A, Derrick G et al. Reaching “An Audience That You Would Never Dream of Speaking To”: Influential Public Health Researchers’ Views on the Role of News Media in Influencing Policy and Public Understanding. Journal of Health Communication 2013;19:260–273. doi:10.1080/10810730.2013.811327

14 Mejia C, Tovani-Palone M, Ticona D et al. The Media and their Informative Role in the Face of the Coronavirus Disease 2019 (COVID-19): Validation of Fear Perception and Magnitude of the Issue (MED-COVID-19). Electronic Journal of General Medicine 2020;17:em239. doi:10.29333/ejgm/7946

15 Lwin M, Lee S, Panchapakesan C et al. Mainstream News Media’s Role in Public Health Communication During Crises: Assessment of Coverage and Correction of COVID-19 Misinformation. Health Communication 2021;:1–9. doi:10.1080/10410236.2021.1937842

16 Chipidza W, Akbaripourdibazar E, Gwanzura T et al. A topic analysis of traditional and social media news coverage of the early COVID-19 pandemic and implications for public health communication. Disaster Medicine and Public Health Preparedness 2021;:1–23. doi:10.1017/dmp.2021.65

17 UK: most popular newspapers 2019 | Statista. Statista. 2021. https://www.statista.com/statistics/246077/reach-of-selected-national-newspapers-in-the-uk/ (accessed 29 Apr 2021).

18 Javadi M, Zarea K. Understanding Thematic Analysis and its Pitfall. Journal of Client Care 2016;1. doi:10.15412/j.jcc.02010107

19 Hewis J. Do MRI Patients Tweet? Thematic Analysis of Patient Tweets About Their MRI Experience. Journal of Medical Imaging and Radiation Sciences 2015;46:396–402. doi:10.1016/j.jmir.2015.08.003

20 Attard A, Coulson N. A thematic analysis of patient communication in Parkinson’s disease online support group discussion forums. Computers in Human Behavior 2012;28:500–506. doi:10.1016/j.chb.2011.10.022

21 Lane J, Hamilton D, MacDonald D et al. Factors that shape the patient’s hospital experience and satisfaction with lower limb arthroplasty: an exploratory thematic analysis. BMJ Open 2016;6:e010871. doi:10.1136/bmjopen-2015-010871

22 Barker K, Subramanian S, Selman R et al. Gender Perspectives on Social Norms Surrounding Teen Pregnancy: A Thematic Analysis of Social Media Data. JMIR Pediatrics and Parenting 2019;2:e13936. doi:10.2196/13936

23 Oyebode O, Ndulue C, Adib A et al. Health, Psychosocial, and Social Issues Emanating From the COVID-19 Pandemic Based on Social Media Comments: Text Mining and Thematic Analysis Approach. JMIR Medical Informatics 2021;9:e22734. doi:10.2196/22734

24 Matza L, Paulus T, Garris C et al. Qualitative Thematic Analysis of Social Media Data to Assess Perceptions of Route of Administration for Antiretroviral Treatment among People Living with HIV. The Patient - Patient-Centered Outcomes Research 2020;13:409–422. doi:10.1007/s40271-020-00417-8

25 Braun V, Clarke V. Using thematic analysis in psychology. Qualitative Research in Psychology 2006;3:77–101. doi:10.1191/1478088706qp063oa

26 Sikdar S, Adali S, Amin M et al. Finding true and credible information on Twitter. 17th International Conference on Information Fusion (FUSION), 2014;1–8.

27 Caulfield T, Clark M, McCormack J et al. Representations of the health value of vitamin D supplementation in newspapers: media content analysis. BMJ Open 2014;4:e006395. doi:10.1136/bmjopen-2014-006395

